# Accurate 3D Positron Range Correction Method for Heterogeneous Material Densities in PET

**DOI:** 10.1101/2022.08.16.22278715

**Authors:** Chong Li, Jürgen Scheins, Lutz Tellmann, Ahlam Issa, Long Wei, N. Jon Shah, Christoph Lerche

## Abstract

**Objective:** The positron range is a fundamental, detector-independent physical limitation to special resolution in positron emission tomography (PET) as it causes a significant blurring of the reconstructed PET images. A major challenge for positron range correction methods is to provide accurate range kernels that inherently incorporate the generally inhomogeneous stopping power, especially at tissue boundaries. In this work, we propose a novel approach to generate accurate three-dimensional (3-D) blurring kernels both in homogenous and heterogeneous media to improve PET spatial resolution.

**Approach:** In the proposed approach, positron energy deposition was approximately tracked along straight paths, depending on the positron stopping power of the underlying material. The positron stopping power was derived from the attenuation coefficient of 511keV gamma photons according to the available PET attenuation maps. Thus, the history of energy deposition is taken into account within the range of kernels. Special emphasis was placed on facilitating the very fast computation of the positron annihilation probability in each voxel.

**Results:** Positron path distributions of ^18^F in low-density polyurethane were in high agreement with Geant4 simulation at an annihilation probability larger than 10^−2^∼10^−3^ of the maximum annihilation probability. The Geant4 simulation was further validated with measured ^18^F depth profiles in these polyurethane phantoms. The tissue boundary of water with cortical bone and lung was correctly modeled. Residual artifacts from the numerical computations were in the range of 1%. The calculated annihilation probability in voxels shows an overall difference of less than 20% compared to the Geant4 simulation.

**Significance:** The proposed method significantly improves spatial resolution for non-standard isotopes by providing accurate range kernels, even in the case of significant tissue inhomogeneities.

## 1. Introduction

Positron emission tomography (PET) is an important, non-invasive *in vivo* medical imaging technique that provides quantitative, molecular information on the distribution of a radiotracer (molecule of interest labeled with a radioactive element) administered to a patient or volunteer (Rahmim *et al*. 2013, Westerterp *et al*. 2007, Avril and Weber 2005). PET imaging works by detecting the annihilation gammas following the positron emission from the radiotracer. However, most of the positrons are thermalized before annihilating; therefore, PET reveals the location of the positron annihilation rather than the location of the emitting nuclide, i.e., the radiotracer. Spatial resolution is a crucial performance parameter in PET imaging, and good spatial resolution is required to enable the precise location of lesions and to quantify uptake in regions of interest (Moses 2011, Sanchez-Crespo *et al*. 2004). The imaging error arising from the positron range is a system-independent physical limitation of the spatial resolution and increases with the positron energy (Cal-Gonzalez *et al*. 2013, Carter *et al*. 2020). This effect is normally neglected in studies applying ^18^F (the most commonly used positron emitter) since it emits positrons with low average energy (approximately 252 keV) and has a maximum positron range of about 2 mm in water (Sahnoun *et al*. 2020, Li *et al*. 2017).

Over the last decade, a number of non-standard positron emitters have been proven to offer significant advantages in specific research topics, for example, ^68^Ga in the diagnosis of prostate cancer and neuroendocrine tumors (Mojtahedi *et al*. 2014, Beheshti *et al*. 2020, Apitzsch *et al*. 2021), ^124^I or ^86^Y in studying slow metabolic processes (Qaim 2011, Cascini *et al*. 2014), and ^82^Rb for cardiac studies. (Chilra *et al*. 2017). Many of these non-standard radioisotopes emit positrons at energies much higher than ^18^F. The average positron energies of ^68^Ga and ^82^Rb are approximately 840keV and 1550keV, respectively, thus leading to a maximum range in water of approximately 9 mm and 17 mm (Li et al. 2017), respectively. Consequently, a significant challenge arises in providing high-quality images when using non-standard positron emitters. Additionally, the continuous development of high spatial resolution PET scanners has increased the demand for fast and robust positron range correction methods. For example, the BrainPET insert for a 7T Magnetic Resonance Imaging (MRI) system currently being built at the Institute of Neuroscience and Medicine, Forschungszentrum Jülich GmbH is expected to achieve a spatial resolution of <2 mm (Lerche *et al*. 2020) over the entire field-of-view (FOV). Modern whole-body scanners, such as the newly developed *Quadra* (Nadig *et al*. 2022) and *Vision* PET-CT scanner from Siemens Healthcare, can also achieve a spatial resolution of close to 3 mm (van Sluis *et al*. 2019, Reddin *et al*. 2018) at the iso-center. The large positron range of non-standard positron emitters, such as ^68^Ga and ^120^I, cause significant degradation of the PET image, an issue which cannot be neglected for such high-resolution PET scanners. Therefore, the development of a fast, accurate, and robust method for positron range correction is an important research avenue.

Strategies for positron range correction in reconstructed PET images have been studied extensively since 1986 (Derenzo 1986), with the majority of approaches using blurring kernels to precisely describe the annihilation distribution. However, the accurate incorporation of structural tissue boundaries into these range kernels is challenging, thus limiting the accuracy of these methods. Although Monte Carlo simulations can obtain accurate range kernels by simulating tissue boundaries in sufficient detail, high-precision or complex simulation-based kernels would be extremely time-consuming (Fu and Qi 2010), making it prohibitive in routine applications. To address this problem, Bai *et al*. (2003 and 2005) developed anisotropic kernels in heterogeneous media by anisotropically truncating Monte Carlo kernels at tissue boundaries or by decomposing the model into a series of paired isotropic convolutions. Alessio *et al*. (2008) have also suggested a rapid estimation method to compensate for the variant kernels by calculating the average fitting parameters of annihilation densities for originating and target voxels. In the studies of Cal-Gonzalez *et al*. (2015), a more realistic kernel was applied in the forward projection step of the iterative reconstruction. Similar to Alessio et al. (2008), Cal-Gonzalez *et al*. (2015) also calculated the average annihilation density but additionally considered all materials surrounding the voxel, thus providing a more reasonable kernel in the case of significant changes in density.

A drawback of the above methods is that they do not correctly consider the path history of the individual positron at tissue boundaries, which inevitably leads to systematic errors. The final annihilation point of the positron depends on its entire energy transfer history, i.e., the tissue types of all voxels passed have an impact on the range via the energy deposition. In this work, we propose an alternative approach in which we use an equation that relates the positron stopping power to the linear attenuation coefficient *μ*_*L*_for 511 keV gamma photons. The deposited energy of the positrons along straight paths through each voxel is tracked in small energy steps compared to the energy deposition in a single voxel. This enables the positron annihilation probability to be computed in every voxel. By correctly taking the path history into account, our proposed method provides a sufficiently accurate range correction for arbitrary tissue boundaries without image segmentation.

## 2. Materials and Methods

### 2.1 Quasi-continuous energy loss (CEL) method

#### 2.1.1 Theoretical model

The positron annihilation probability along the trajectory is computed based on the energy loss process of the positron, which can be expressed using the positron stopping power (Berger and Seltzer 1983, Kheradmand Saadi and Machrafi 2020) as:

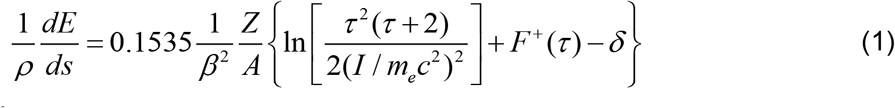

with 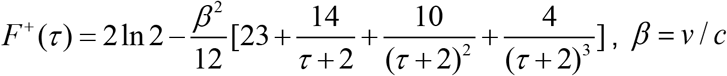 and *τ* = *E* / *m*_*e*_*c*^2^. Here, *ρ* is the absorber density, *v* and *c* are the velocities of incident positron and light, Z and A are the atomic number and atomic weight, *E* is the kinetic energy of incoming positron, *m*_*e*_*c*^2^ is the electron rest energy, *I* is the mean excitation energy of the target, and *δ* is the density-effect correction. The path length through which a positron of any energy can pass, i.e. the path length table, can be approximated (Kheradmand Saadi and Machrafi 2020) with

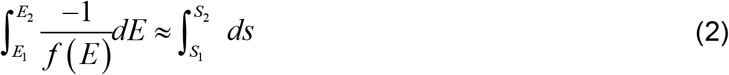

where 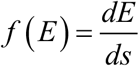 is the positron linear stopping power, and *E*_*1*_ and *E*_*2*_ are the positron energy at path lengths *S*_*1*_ and *S*_*2*_. Using the path length table and positron initial energy sampled from the energy spectra of positron emitters, the remaining energy of the positron passing through each voxel is calculated to determine whether the positron can leave the current voxel. Accordingly, the positron annihilates in the current voxel or moves into the next one.

To build a tissue-dependent range kernel, tissue information is incorporated in the calculation of the energy deposition according to eq. (1), but is dependent on the attenuation coefficient of the 511 keV gamma photons. The attenuation coefficient is required for attenuation correction during PET reconstruction (Kinahan *et al*. 1998) and is, therefore, always available via the attenuation map for all image voxels. Thus, it allows us to implement the tissue dependence of each voxel via its assigned value (Kops *et al*. 2015). The linear attenuation coefficient *μ*_*L*_ can be expressed as eq. (3) (Arslan 2019):

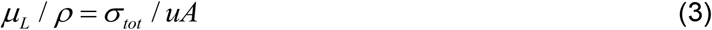

with the total cross section written as: *σ* _*tot*_ = *σ* _*ph*_ + *σ* _*coh*_ + *σ*_*incoh*_ + *σ* _*pair*_ where *u* is the atomic mass unit, *σ* _*ph*_is the cross section of atomic photo-effect, *σ* _*coh*_and *σ* _*incoh*_ are the Rayleigh and the Compton scattering cross sections, and *σ* _*pair*_is the cross sections for electron-positron production. For typical tissues and gamma energies close to 511 keV, the cross section *σ* _*incoh*_of Compton scattering dominates all other elementary processes by at least two orders of magnitude (Berger *et al*. 2010). The Compton scattering cross section per atom can be written as the product of the scattering cross section per electron *σ* _*e*−*incoh*_and the atomic number Z. Consequently, the linear attenuation coefficient can be approximated as:

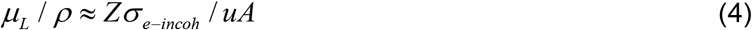

Here, *σ* _*e*−*incoh*_ is the electronic Compton scattering cross sections. By replacing the item *Z* / *A* in eq. (1) and (4), the linear attenuation coefficient *μ*_*L*_ is incorporated into the linear stopping power eq. as follows:

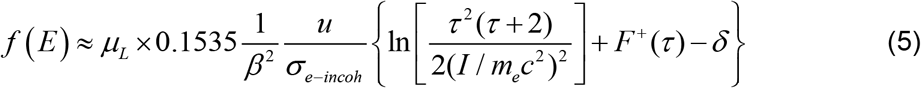

The *μ*_*L*_ can be replaced with the product of the attenuation coefficient in water *μ*_*Water*_ and the relative attenuation coefficient of other media to water *μ*_*R*_. Consequently, the tissue-dependent linear stopping power is further simplified as *f* (*E*) ≈ *μ*_*R*_ × *f* (*E*)_*water*_, where *f* (*E*)_*water*_ is the positron linear stopping power in water. The tissue-dependent path length table can be approximated as:

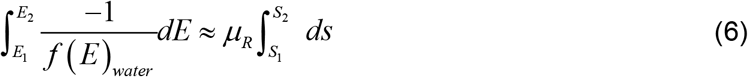

The annihilation probability in the 3-D space was computed by applying the mathematical model in all directions. The distribution of annihilation probability herein referenced the total positron path length rather than the positron range (straight length between source and annihilation point). The repeated collisions with the electrons lead to an effective range that is always shorter than the total path length. To take this effect into account, we approximated the reduction of the positron range compared to the total path length by multiplying the stopping power by a correction factor larger than 1. The correction factor depends on the positron initial energy. The correction factor was obtained by fitting an empirical model to the positron ranges in water for positron energy ranging from 10 keV to 5000 keV, as in eq. (7) where *e* is the Euler number and *E* is the positron energy.

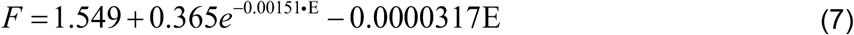

#### 2.1.2 Path segment length in each voxel

By using eq. (6), we only need the stopping power in water, the attenuation map, and the path segment lengths of the positron trajectory through each voxel to calculate the annihilation probability in all voxels. The path segment lengths vary according to the directions and position of positrons entering the voxel (see the red track in Fig. 1). To calculate the path segment length in successive voxels, an annihilation ball was constructed by discretizing the sphere into a sufficiently large number of similar triangular cells (Fig. 1). For a discretization with sufficiently small triangles, the line connecting the center of the triangle to the center of the sphere is a good approximation for all possible positron paths inside the polyhedron spanned by the sphere’s center and the surface triangle. Based on this model, the path length was calculated by recording the intersection of the red path with each voxel boundary (see the red intersection in Fig. 1). By applying this method for all directions, a look-up table (LUT) was created in which the path ID, the voxel ID, and all path segments were stored.

**Figure 1.**
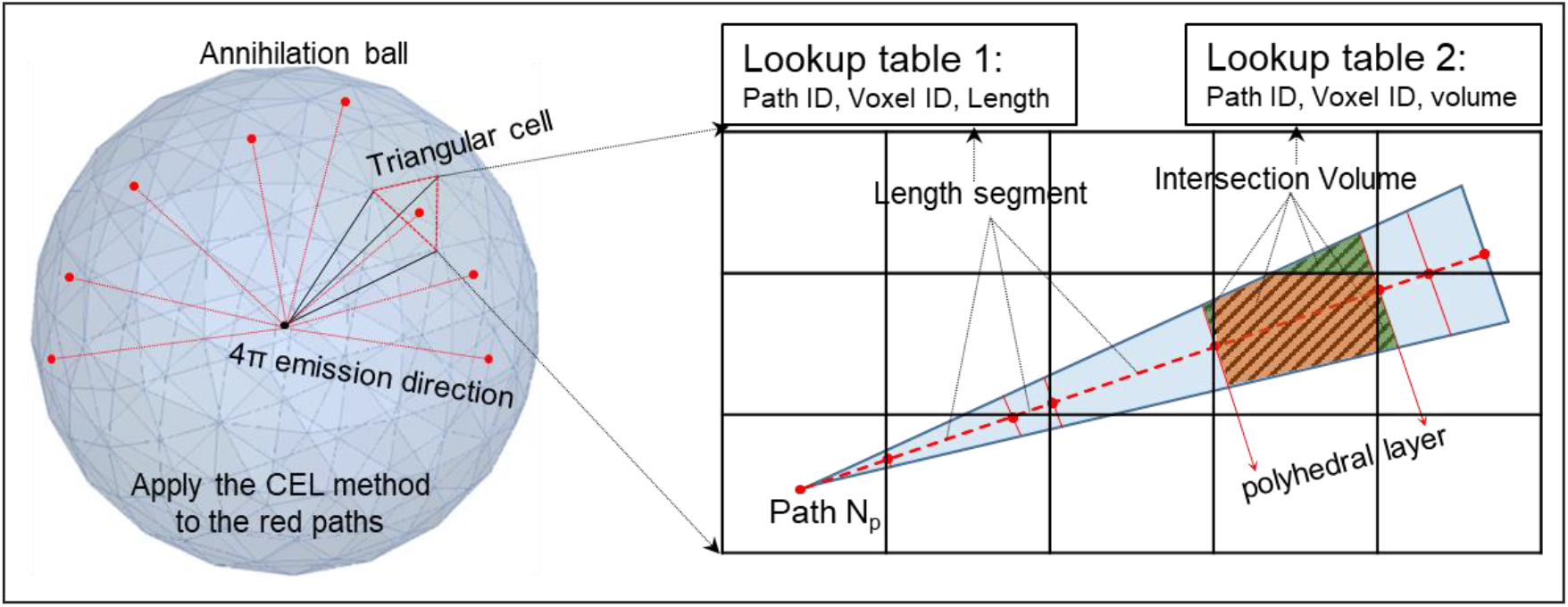
The schematic diagram for calculating the positron path length and the volume of interacting parts in each voxel

#### 2.1.3 Annihilation probability contribution for each voxel

The calculated annihilation probability approximately spreads uniformly over the polyhedral layers corresponding to each line segment (grey shaded area in Fig. 1). Thus, the contribution of all polyhedral layers to the annihilation probability of the surrounbding voxels can be approximated using the intersecting volume of the corresponding polyhedral layer and the voxel (green and orange areas in Fig. 1). In this case, the annihilation probability contribution for the voxel can be calculated according to the volume ratio according to

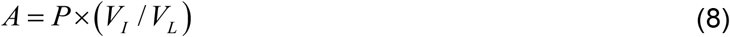

where P is the cumulated positron annihilation probability along each path segment. *V*_*I*_ / *V*_*L*_ is used to calculate the annihilation probability assigned to each intersection volume, *V*_*I*_(green and orange areas), and *V*_*L*_is the volume of each polyhedral layer associated with the path segment (grey shaded area). A LUT of the *V*_*I*_ along with the path ID and voxel ID was precomputed using the model in Fig. 1.

With respect to every positron path ID and voxel ID, the path segment length and interacting volumes in each voxel were obtained from the LUTs in Fig. 2, which were created based on the geometric models in Fig. 1. In order to assure the consideration of all significant annihilation probability contributions, a sufficiently large array of 33*33*33 voxels with a voxel size of 1.25*1.25*1.25 mm was created. This array size corresponded to an annihilation ball with a 20 mm radius – large enough for all studied isotopes and tissue types, except for the combination of ^120^I and lung tissue, which was omitted at the current stage due to the high computational burden required for creating the LUTs. A total of 5120 straight, radial paths, homogeneously distributed over the sphere, were considered in the computation. The algorithm of the theoretical model is given in the appendix.

**Figure 2.**
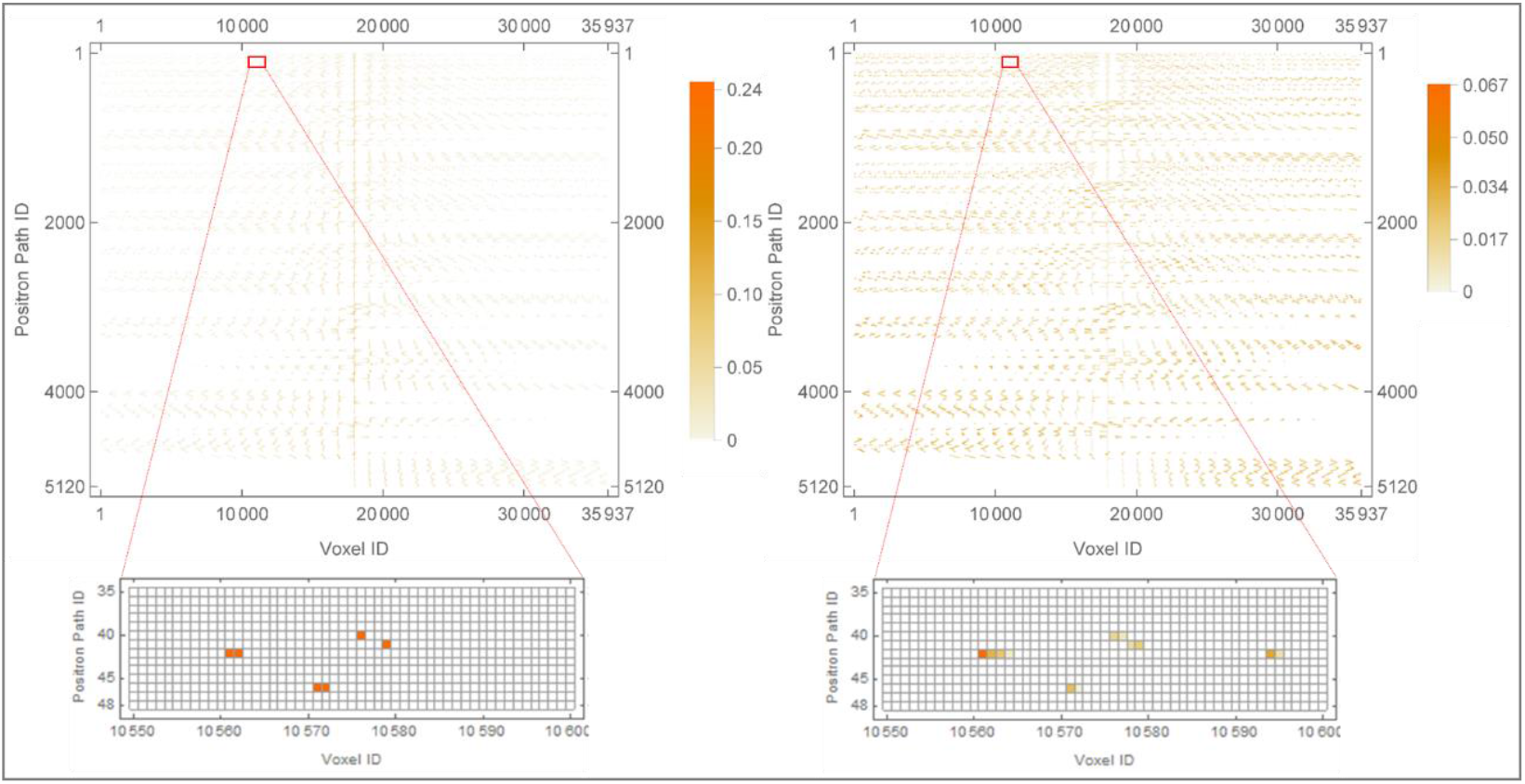
Look-up tables of the path segment length and interacting volumes with respect to each path ID and voxel ID.

### 2.2 Phantom experiments

The phantom experiments were carried out with using an aqueous solution of ^18^F in the hybrid 3T MR-BrainPET operated in Forschungszentrum Jülich (Fig. 3 (c)) (Caldeira *et al*. 2019). The phantoms were used to compare the measurement with the Geant4 model against our method, tested in 3-D space. Due to the low positron energy of ^18^F, very low-density polyurethane blocks were used to enable measurable depth profiles. The densities of the materials were 0.01 g/cm^3^, 0.016 g/cm^3^, 0.035 g/cm^3^, and 0.089 g/cm^3^, respectively. In these polymers, positrons emitted from ^18^F could travel a maximum distance of 150 mm. A planar, disk-shaped ^18^F source with a diameter of 4 cm was placed at the center of the surface of every test polymer block (200 mm cube). The scanner covers an axial FOV of 19.2 cm and a transaxial FOV of 31.4 cm in diameter. A constant 3 T magnetic field without coils was employed in this experiment, leading to a strong shortening of the positron range in the transaxial direction. Consequently, almost no positrons escape from the blocks due to the additional compression forcing the positrons on tight spiral trajectories in the axial direction. A layered block constructed from a combination of the aforementioned materials and foams in the following order: #1(10 mm)-#4(10 mm)-#1(20 mm)-#4(base) was also designed to verify the positron depth profiles in heterogeneous materials. The image was reconstructed using a vendor-provided ordinary Poisson ordered subset expectation maximization (OP-OSEM) method with two subsets and 32 iterations (Lerche *et al*. 2021). No scattering or attenuation correction was employed during the image reconstruction as no gamma scattering or attenuation was expected for these low-density materials. The annihilation probability of voxels along the axial FOV was obtained from the reconstructed images consisting of 256 × 256 × 153 isotropic voxels of 1.25.

**Figure 3.**
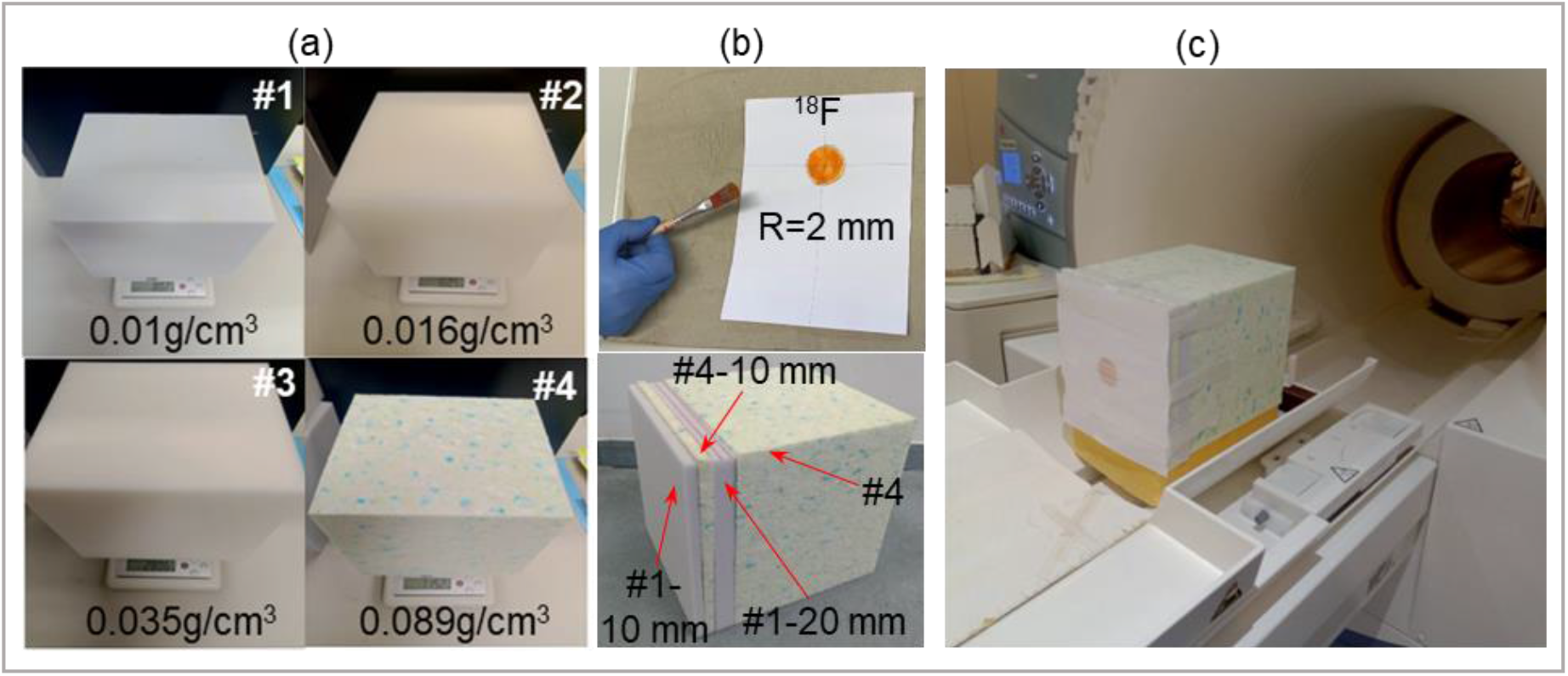
(a) Low-density polymers block for measuring the ^18^F implantation profiles. (b) 4 cm diameter ^18^F disk source painted on paper and the structure of layered polymers block. (c) Setup for measuring the implantation profiles in layered polymers block.

### 2.3 Geant4 simulation

In this work, the *G4EmStandardPhysics* model was used to simulate the electromagnetic processes of positron and gamma transport through matter. The Geant4 toolkit was used for the Geant4 simulations applied in this study. The toolkit is designed to simulate the interaction of particles through matter, with medical physics being an important area of application (Allison *et al*. 2006).

The Geant4 toolkit was used to simulate both the stopping power of positrons in water and the energy spectra of selected positron emitters, thus enabling the creation of LUTs. The toolkit was additionally used for verifying positron range and path length in one-dimensional (1-D) space and the positron range induced point spread functions (PSF_PR_) in 3-D space. For the verification of path length, the structure and composition of the phantom measurement were reproduced in the Geant4 simulations. With respect to the verification of the PSF_PR_ functions, the point sources of ^18^F, ^68^Ga, and ^120^I were placed at the center of a cube filled with different materials and tissues. i.e., water, cortical bone, lung, and mixtures. In the volumes with mixed *μ*_*R*_ values, the voxels from ID-1 to ID-17 were filled with values representing water for ^120^I and lung tissue for ^18^F, and the voxels from ID-18 to ID-33 were filled with values representing cortical bone for ^120^I and water for ^18^F. The composition of the tissues was taken from the National Institute of Standards and Technology (NIST) material database (available online at: https://www.nist.gov/pml/x-ray-mass-attenuation-coefficients).

Finally, the setup in Fig. 4 was simulated to record the intensity of the 511 keV gamma photons hitting the detector without attenuation and scattering. Samples of the four polymers, in thicknesses of 200 mm, 150 mm, 100 mm, and 70 mm, were placed between two 100 mm thick lead collimators. The attenuation coefficient was calculated *μ*_*L*_ *x* / *ρ* = ln (*I*_0_ / *I*), where *I*_*0*_ and *I* are the incident and attenuated photon intensity, *ρ* is the density, and *x* is the sample thickness. The Geant4 model shows a high precision with relative errors of less than 0.4% in water. The simulated relative attenuation coefficient *μ*_*R*_ of the four polymers were 0.01, 0.016, 0.035, and 0.088 in order of increasing density.

**Figure 4.**
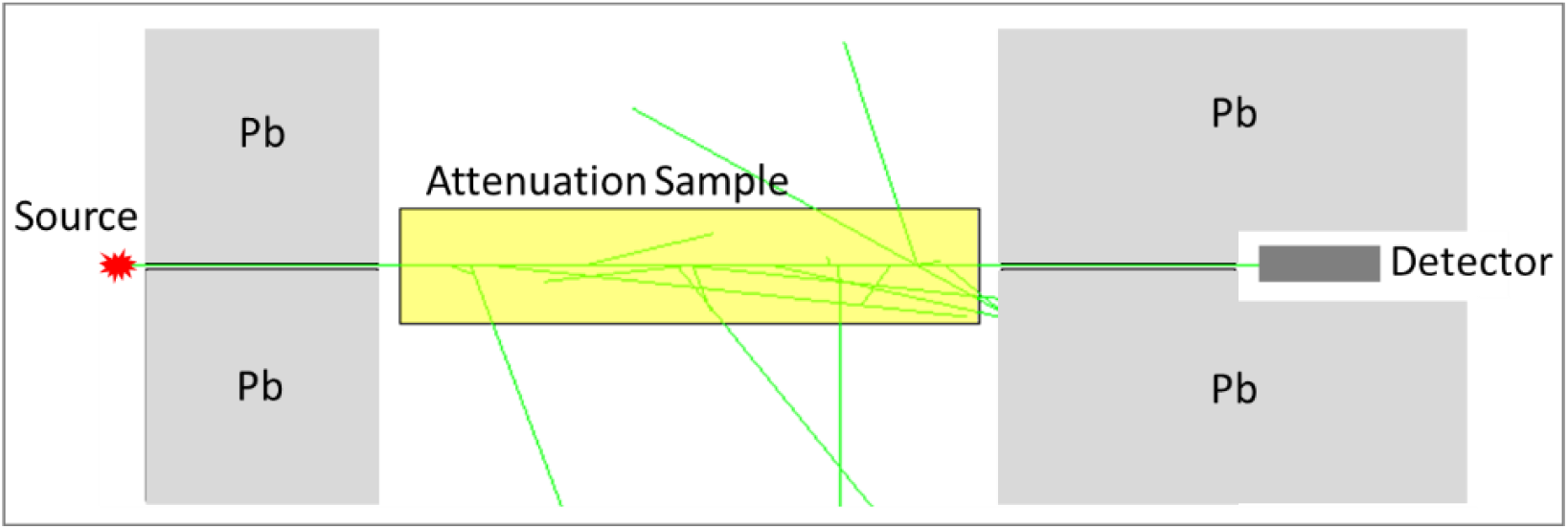
Schematic diagram of the Geant4 setup for simulating attenuation coefficient.

## 3. Results

### 3.1 Verification of the theoretical model

The positron path length in media different to water is approximately proportional to the relative attenuation coefficient *μ*_*R*_with respect to eq. (6). The theoretical model was validated using a Geant4 simulated path length in water, B-100 bone-equivalent plastic, C-552 air-equivalent plastic, and cortical bone, as shown in Fig. 5. The *μ*_*R*_ of above mentioned media, which are 1, 1.381, 1.585, and 1.788, respectively, covers the range of most human tissue types. The plots of Fig. 5 were obtained by scaling the path lengths in the three media to water. This was achieved by multiplying the x-value and dividing the y-value with the relative attenuation coefficient *μ*_*R*_ values. The scaled curves show very good agreement with the simulated curves in water with root mean square deviations (RMSD) of 0.0015, 0.0021, and 0.0026, respectively.

**Figure 5.**
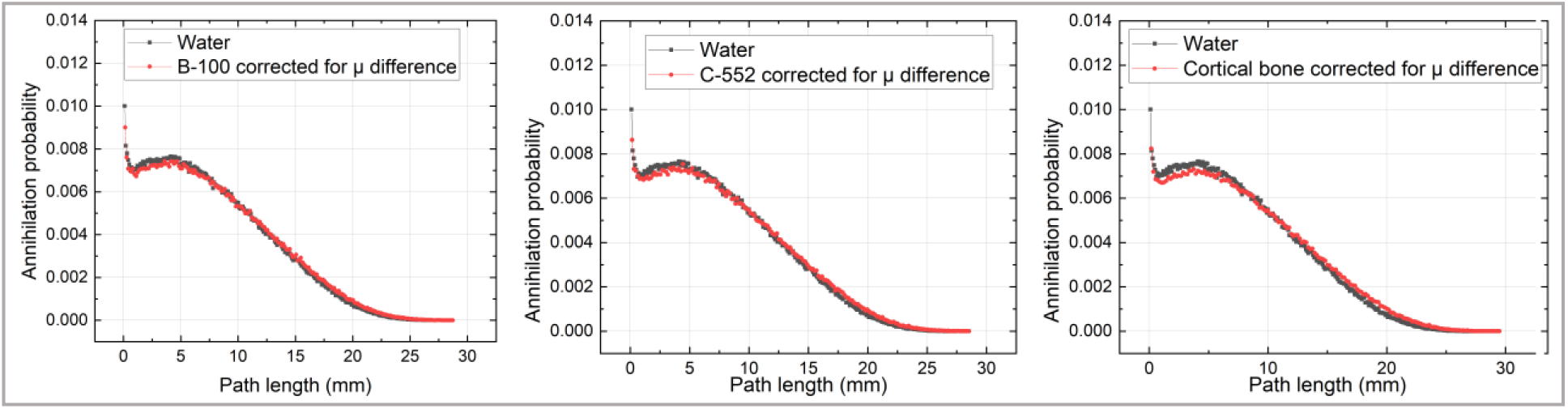
Comparison of simulated positron path length in water with *µ*_*R*_-scaled positron path lengths in B-100 bone-equivalent plastic, C-552 air-equivalent plastic, and cortical bone

### 3.2 Energy spectra and positron stopping powers of positron emitters

Fig. 6 shows the simulated energy spectra of the positron emitters ^18^F, ^68^Ga, and ^120^I. The maximum and average positron energy of the positron emitters are 0.231/0.585 MeV, 0.76/1.865 MeV, and 1.59/4.495 MeV, respectively. In this context, the results of the positron emitters can illustrate the model’s accuracy for most typical positron emitters (Qaim 2011).

**Figure 6.**
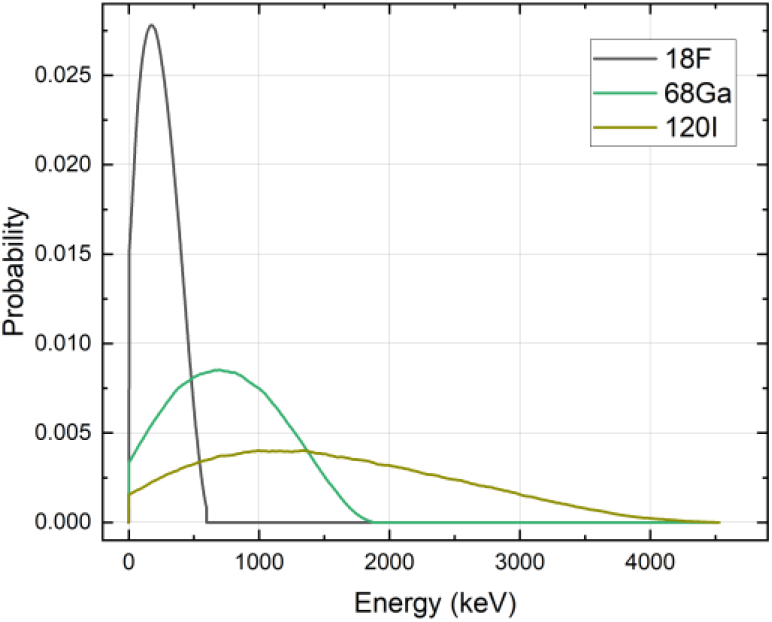
Energy spectra of selected positron emitters were simulated using the Geant4 toolkit.

Fig. 7(a) shows the stopping power of positrons in water with and without stopping power correction according to eq. (7). The energy step for the tables is 1 keV, and the values in between are linearly interpolated. With the corrected stopping power, the approximated range of monoenergetic positrons in water also matches the simulated average range shown in Fig. 7(b). Fig. 7(c) shows the ratio of path length to range together with the best fit (Eq. (7)) of the parametrization for the correction factors. Table 2 shows the best-fit parameters, the fit parameter errors, and the p-values. Range corrections for materials other than water are derived from the range table shown in Fig. 7(b) by multiplication with the relative *μ*_*R*_ value.

**Table 2.** The best-fit parameters, the fit parameter errors, and the p-values of eq. (7)

|  | Estimate | Standard Error | P-Value |
| --- | --- | --- | --- |
| $A$ | 0.365 | 0.0462023 | $7.11085 \cdot 10^{-11}$ |
| $\lambda$ | 0.00151 | 0.0003942 | 0.000313945 |
| $c$ | -0.0000317 | 0.0000127 | 0.0151906 |
| $d$ | 1.549 | 0.0488827 | $3.54903 \cdot 10^{-39}$ |

**Figure 7.**
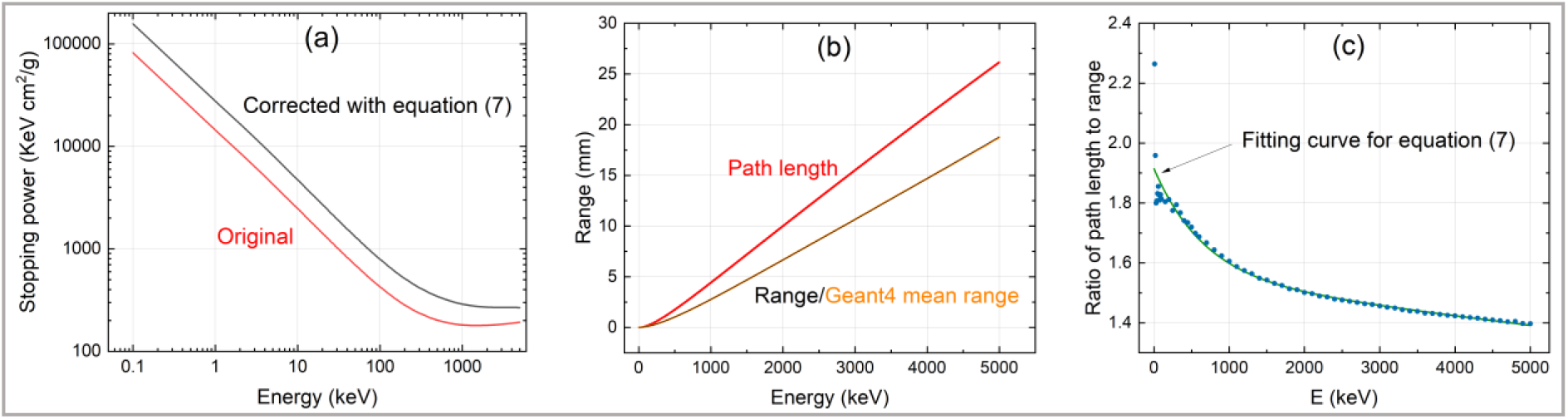
(a) positron stopping power in water without and with exponential correction. (b) the range of monoenergetic positron in water without and with exponential correction. (c) the ratio of path length to range and the fitting curves of eq. (7).

### 3.3 Positron path length in 1-D space

The path length for a 1-D space example was computed with the CEL model and was verified against the Geant4 simulation. The path length of ^18^F in blocks #1 and #4 is in very good agreement with the Geant4 simulation at annihilation probability larger than 10^−2^∼10^−3^ of the maximum annihilation probability, as shown in Fig. 8. The RMSD between the simulated and measured values for blocks #1 and #4 were 0.0363 and 0.0049, respectively, when normalizing the annihilation probability profiles to the maximum value. The relative differences in annihilation probabilities were smaller than 10% for the majority of path lengths. Relative differences exceeded 30% at annihilation probabilities smaller than 10^−2^∼10^−3^ of the maximum annihilation probability, which would not affect the correction, as these values are excluded by a lower threshold for annihilation probabilities in a single voxel. The optimum value for this low threshold still needs to be determined in an independent study. Appropriate values are in the range of 1% to 10%, as the quantification accuracy of PET is currently at this level.

**Figure 8.**
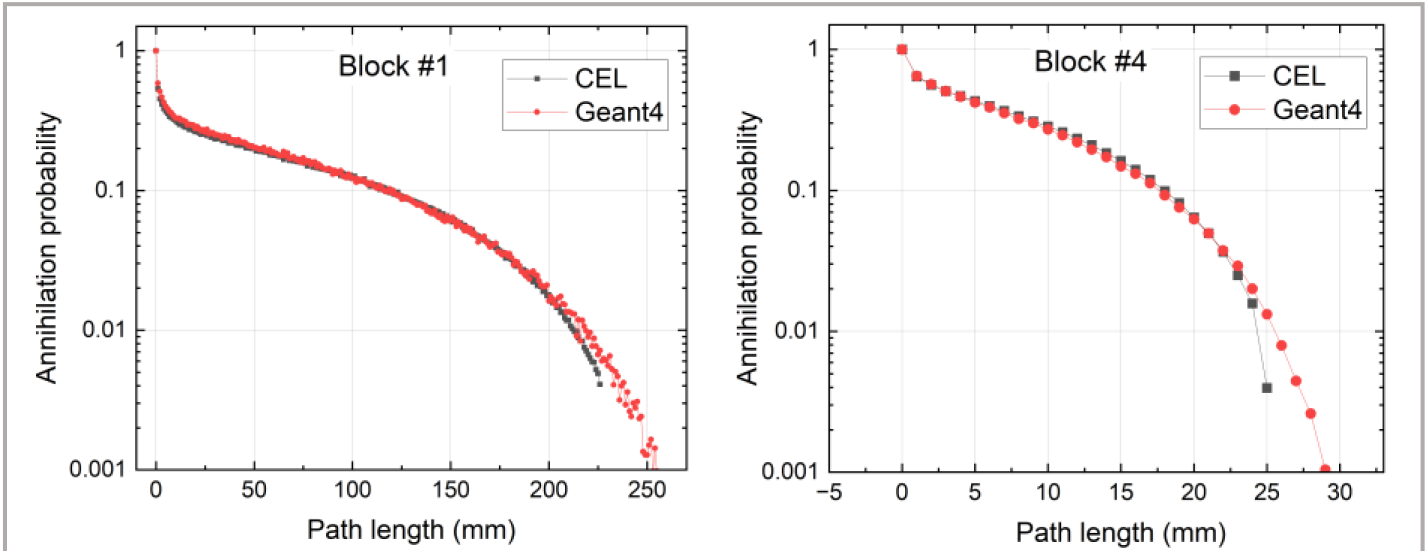
Geant4 simulated (red) and CEL method generated (black) path length of ^18^F in block #1 and block #4.

The positron depth profiles simulated using Geant4 in single and layered blocks were verified against measured profiles obtained with the 3T MR-BrainPET scanner, as shown in Fig. 9. It can be seen that the simulated and measured profiles in single blocks are in very good agreement. The calculated RMSD were 0.0531, 0.0058, 0.081, and 0.008, respectively, when normalizing them to the maximum annihilation probability. The simulated profiles for the layered block were convolved with a Gaussian distribution of 3 mm full width at half maximum (FWHM) according to the image resolution of the 3T MR-BrainPET scanner close to the center. The layered phantom leads to an irregular annihilation probability distribution, especially at the media boundaries. In this case, simulations and measurements are also in good agreement. The observed difference can be explained by potential air enclosures and the inhomogeneous density of the #4 block, which are present in the measurement but have not been included in the Geant4 simulation. As a consequence, we modified the density of slice #4-10 mm for the simulation and set it to 0.69 g/cm^3^ instead of 0.89 g/cm^3^. By doing this, we obtained an annihilation probability profile that was in better agreement with the measurements, and the corresponding RMSD dropped from 0.0168 to 0.0004.

**Figure 9.**
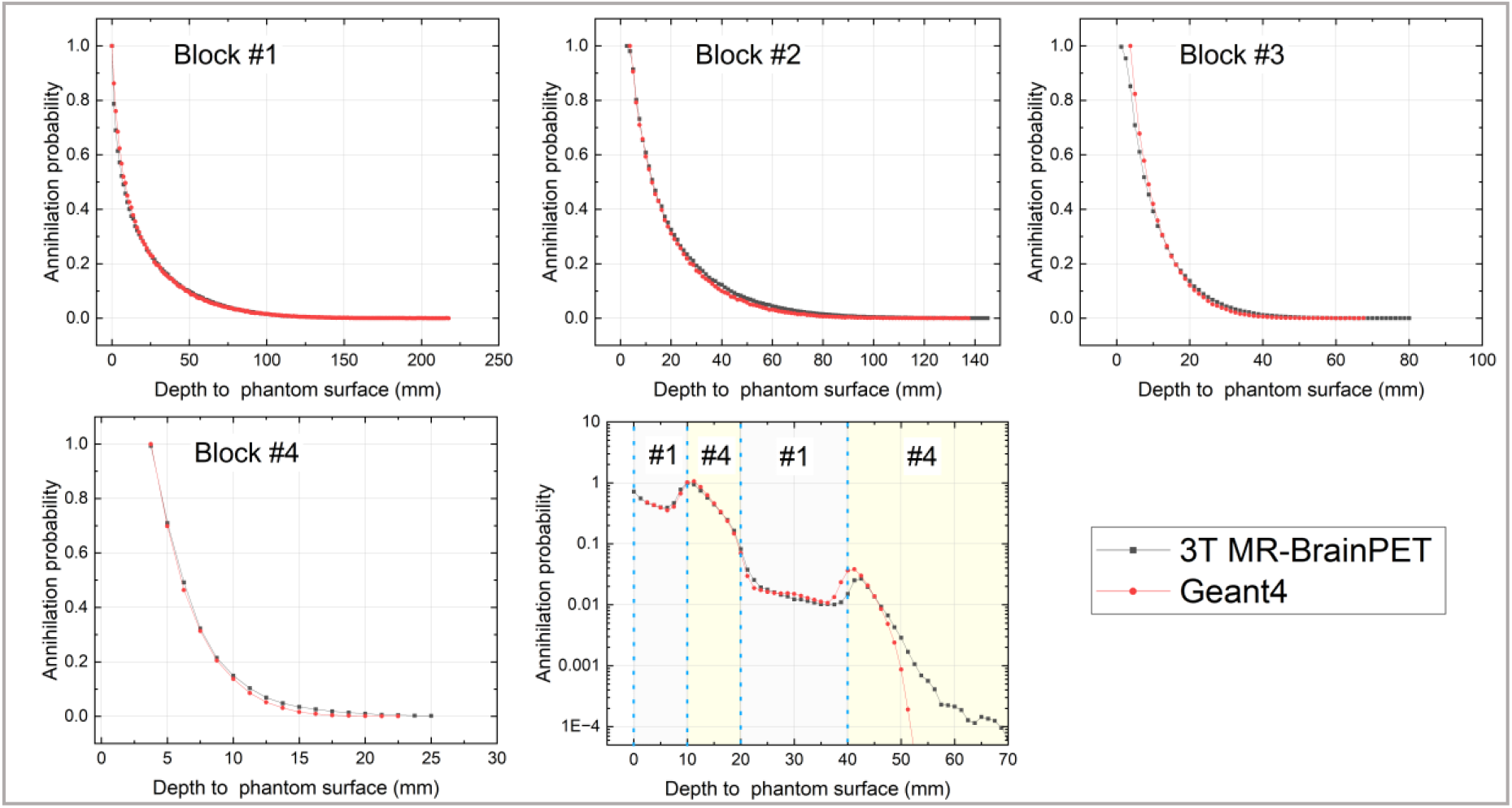
Geant4 simulated (red) and measured (black) profiles of positron annihilation probability versus axial depth in single-layered and layered blocks.

### 3.4 PSF_PR_

Fig. 10 shows the Geant4 simulated and CEL computed annihilation probability distribution of _120_I and ^18^F in water, cortical bone, lung tissue (the *μ*_*R*_ is 0.1213), and the volumes with mixed *μ*_*R*_ values. In the case of significant tissue changes from water to the bone and lung, the tissue boundaries were modeled correctly according to the tissue distribution simulated using Geant4. Fig. 11 shows the simulated and CEL-generated PSF_PR_ of ^18^F, ^68^Ga, and ^120^I for the above-mentioned tissue mixtures at a centric slice of the 3-D voxel array. The annihilation probability in the voxels at the centric slice was normalized to the overall annihilation probability of the entire 3-D voxel array to obtain the final PSF_PR_ functions. By applying the corrected stopping power, the relative differences of annihilation probabilities did not exceed 20% for those voxels with an impact on spatial resolution (the ones surrounding the central voxel of the PSF_PR_ functions with annihilation probabilities larger than the 10^−2^∼10^−3^ fold of the maximum annihilation probability). Although Figs 10 and 11 show large relative differences in annihilation probabilities for the voxels at annihilation probabilities smaller than 10^−3^ of the maximum annihilation probability, these can be neglected owing to the explanations outlined in section 3.3. An exception is the relative difference of ^18^F in the first water-filled voxel, which increases to 50%. However, this can also be ignored as the small range of ^18^F in water resulted in no positron escape from the first voxel.

**Figure 10.**
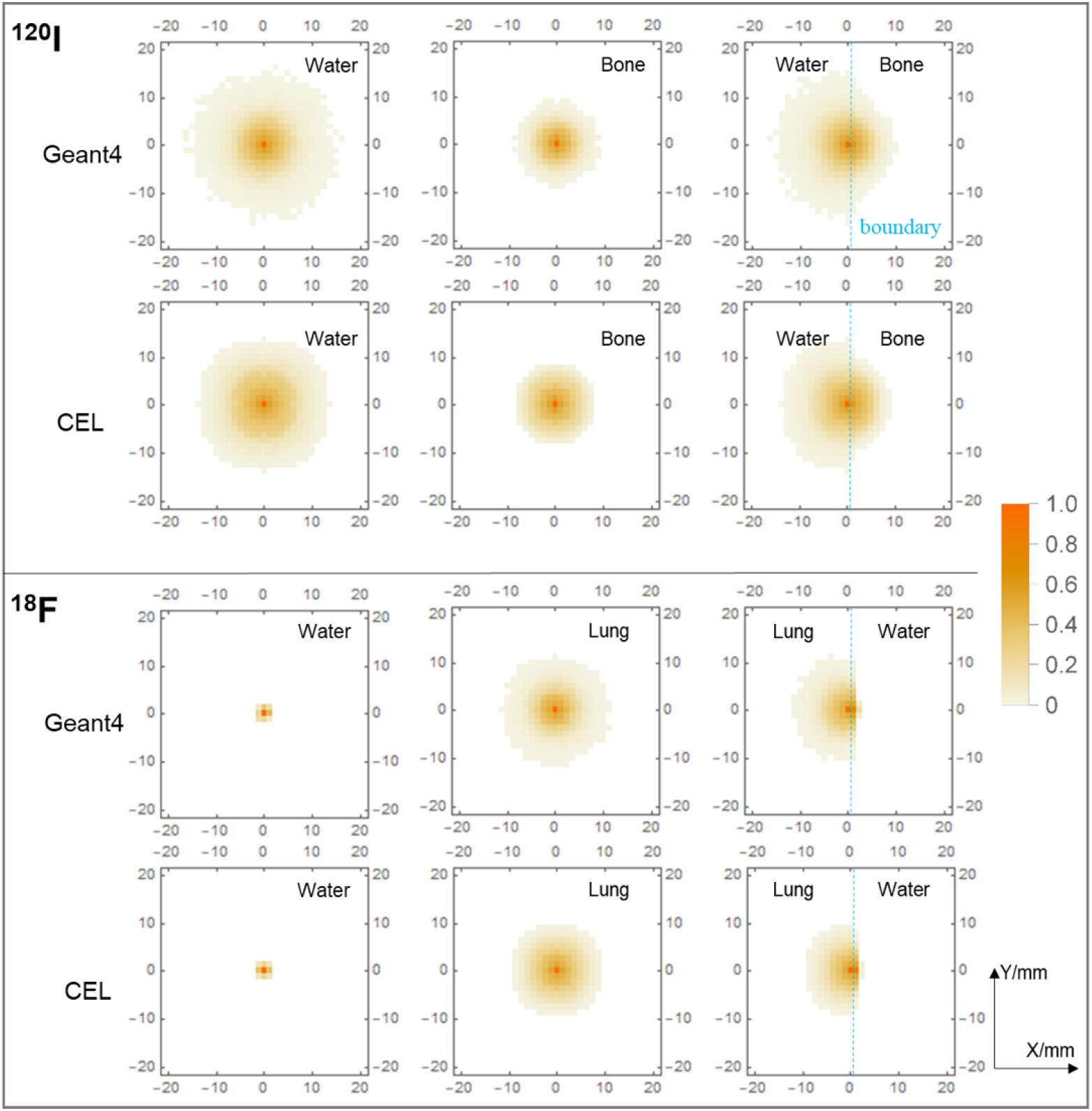
Positron range induced annihilation probabilities of ^120^I and ^18^F in water, cortical bone, lung, and the volumes with mixed *μ*_*R*_ values computed with Geant4 and CEL method. The lower threshold for annihilation probabilities in a single voxel was set to 10^−4^ of the maximum annihilation probability.

**Figure 11.**
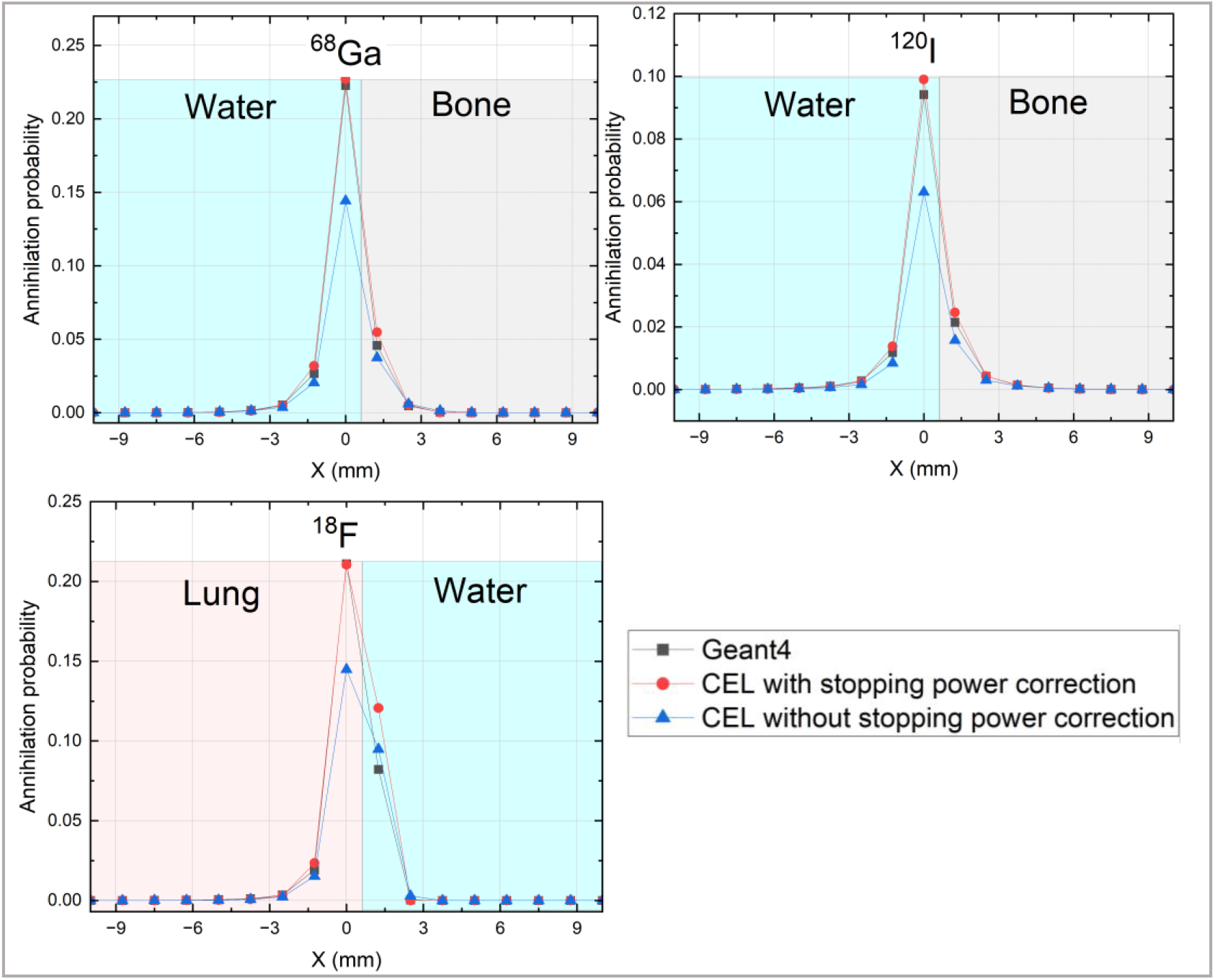
PSF_PR_ of ^18^F, ^68^Ga, and ^120^I in mixtures of lung/water and water/cortical bone. The central voxel (edges from −0.625 mm to 0.625 mm) was filled with lung for ^18^F and water for ^68^Ga and ^120^I, respectively.

## 4. Discussion

An important feature of the proposed CEL method is that the positron range is calculated based on its energy deposition, step by step, in each voxel. Consequently, the energy deposition across tissue boundaries can be easily calculated voxel by voxel by considering the corresponding attenuation coefficient value, which is available through the attenuation map. No additional image segmentation (Kertesz *et al*. 2022) (Cal-Gonzalez *et al*. 2015), assignment of tissue types, or average kernel approximation at the boundary (Cal-Gonzalez *et al*. 2015, Alessio *et al*. 2008)(Kraus *et al*. 2012) is required for computing the positron distribution in heterogeneous media. With our approach, the positron range-induced PSF_PR_ functions take the accumulative energy loss along the entire positron path into account and correctly consider boundaries in heterogeneous media. Consequently, compared to the latest work of Kertesz *et al*. (2022), in which an attenuation map was also used to obtain PSF_PR_ functions in heterogeneous media, our method is expected to be more robust and better at modeling the boundaries of different tissues. For example, in the case of a positron traveling from lower to higher density material, our method presents a much lower deviation compared to simulations (Fig. 10 and 11) when calculating the positron annihilation probability at tissue boundaries. This is because our method only requires the corresponding and known attenuation coefficient, thus excluding potentially erroneous segmentation. In addition, as most of the required time-consuming computations are object independent, they can be conducted in advance and stored in LUTs. Only the step-wise computation of the energy loss against the straight positron paths is required to generate the range kernel for the reconstruction, which takes less than 10 minutes at the current kernel size. Furthermore, the PSF_PR_ functions depend only on the attenuation map, not on the emission image, and only need to be computed once during the image reconstruction.

The proposed CEL method was validated in three steps. First, the theoretical approach was validated using Monte Carlo simulation with Geant4. Here, a good agreement with the results was observed. A fundamental feature of our CEL approach is that, by using a *µ*_*R*_ value relative to water when computing the energy deposit, the approximations of the positron annihilation probabilities for different tissue types are based on those of water. It is shown that the approximation generates well-matched path lengths for different tissues when compared to those generated by Geant4 for water. The accuracy of this approximation is sufficient for calculating the tissue-dependent positron annihilation probabilities to be used for positron range correction in PET. Thus, the stopping power for any tissue and voxel can be easily calculated by using the corresponding attenuation coefficient of 511 keV gamma photons, which is readily available via the attenuation map required for attenuation corrections.

Having validated theoretical model, the method was then verified with Monte Carlo simulation and phantom experiments by computing the path length of ^18^F in very low-density polyurethane blocks in 1-D space. Geant4 simulation is introduced herein as an intermediate reference to cross-validate the CEL method with the phantom experiment due to the difficulties in directly obtaining path length distribution for model validation using the scanner. The very low density of the polymers resulted in a wide distribution of the maximum path length of ^18^F from 25 mm to 250 mm and enabling the positron depth profiles to be measured in the axial direction with the Siemens 3T BrainPET insert, despite its limited spatial resolution of ≈ 3 mm at the center. In this context, the well-matched profiles indicate the validity of the proposed method for different typical tissue *µ*_*R*_ values.

Finally, the proposed method was applied to 3-D space for computing the annihilation probability in each voxel and for generating the positron range-induced PSF_PR_ functions. The results were verified against the Monte Carlo simulation with Geant4. Exemplarily cortical bone and lung tissue values were used for this validation as they are the tissues in the human body with the highest and lowest densities, respectively. The 2-D distribution of the positron annihilation probability in each voxel, as shown in Fig. 10, demonstrates the validity of the CEL method in modeling tissue boundaries, even in the case of significant changes in the attenuation coefficient. The 1-D profile of the estimated PSF_PR_ functions of ^18^F, ^68^Ga, and ^120^I for tissue mixtures are also shown in Fig. 11. Again, good agreement between Geant4 and CEL can be observed. We have therefore shown that the CEL method is suitable for effectively computing the tissue-dependent PSF_PR_ functions required for improving spatial resolution for typical standard or non-standard positron emitters in PET imaging. As the correct prediction of the PSF_PR_ is of high importance for PET image de-blurring, we expect a significant improvement in PET spatial resolution, especially for nuclides with high positron energy (Shah *et al*. 2014).

The accuracy of the CEL model is highly dependent on the accuracy of the LUTs of path length and interacting volume in each voxel. To enable fast calculation, two reasonable approximations were made. First, for the computation of the energy deposit, the positrons are considered to fly along straight lines connecting the incident and outgoing positions in each voxel. Second, the annihilation probabilities computed for this straight path (red path segment in Fig. 1) are used for the entire tetrahedron. Differences arise from the discrepancy between the complex real paths and the straight paths shown in Fig. 1 when calculating the annihilation probabilities. The LUTs depend on the specific discretization of the sphere and the sizes and positions of the image voxels, but not on the attenuation coefficients. The shift-invariant LUTs are, therefore, object-independent and can be precomputed and applied to all objects. In addition, the calculation only depends on the geometric setting. A re-computation is only required if the voxel size or the discretization of the sphere is changed. The differences between the Geant4 simulations and the measurements can be further reduced by considering more positron paths, i.e., using a more fine-grained discretization of the sphere. However, this will significantly increase the computational effort.

## 5. Conclusion and further works

This study proposes a novel and robust method to compute the positron range-induced PSF_PR_ functions. By using an explicit formula that relates the positron stopping power to the linear attenuation coefficient for 511 keV gamma photons, this method considers the positron path history in step sizes much smaller than the typical voxels size and allows for sufficiently fast computation due to the precomputed LUTs, making it suitable for clinical applications. The calculated path lengths, tissue boundaries, and the PSF_PR_ functions of ^18^F, ^68^Ga, and ^120^I were all in good agreement with those obtained using Monte Carlo simulations. Phantom measurements were used to verify the accuracy of the simulated positron annihilation profiles for a directed positron beam in a magnetic field. Based on the approximations used, the relative systematic error of the positron annihilation in one voxel was smaller than 20%. The boundaries between tissues with high differences in attenuation coefficient, such as water to cortical bone and water to lung tissue, were reproduced correctly using the proposed method. Thus, the novel method is robust and is suitable for standard or non-standard positron emitters with comparable positron energies and for all typical cases of tissue compositions.

The anisotropy of the PSF_PR_ is not only caused by inhomogeneous media and may also be affected by the presence of magnetic fields in PET/ magnetic resonance imaging (MRI) hybrid scanner systems. The Lorentz force only compresses the transverse positron annihilation distribution, so the transaxial distribution might be overcorrected if a homogeneous kernel is applied. In this case, a magnetic field-based anisotropic range kernel is required. Thus, future work will focus on extending the method to make it compatible with combined PET/MRI imaging through the application of a correction to the CEL method in the presence of a magnetic field.

## Data Availability

All data produced in the present study are available upon reasonable request to the authors

## Acknowledgments

The research was supported by the Helmholtz-OCPC (Office of China Postdoc Council) Postdoc-Program through the Forschungszentrum Jülich GmbH. This work was also supported by the China National Natural Science Foundation (Grant Nos. U1932162). The authors thank Ms. Claire Rick for English proofreading and Mr. Markus Lang for the preparation of the F18-solution.

## Appendix Pseudocode

A pseudo-code description of the algorithm used to compute the 3-D positron annihilation probability in each voxel.

### Algorithm: Quasi-continuous energy loss (CEL) method

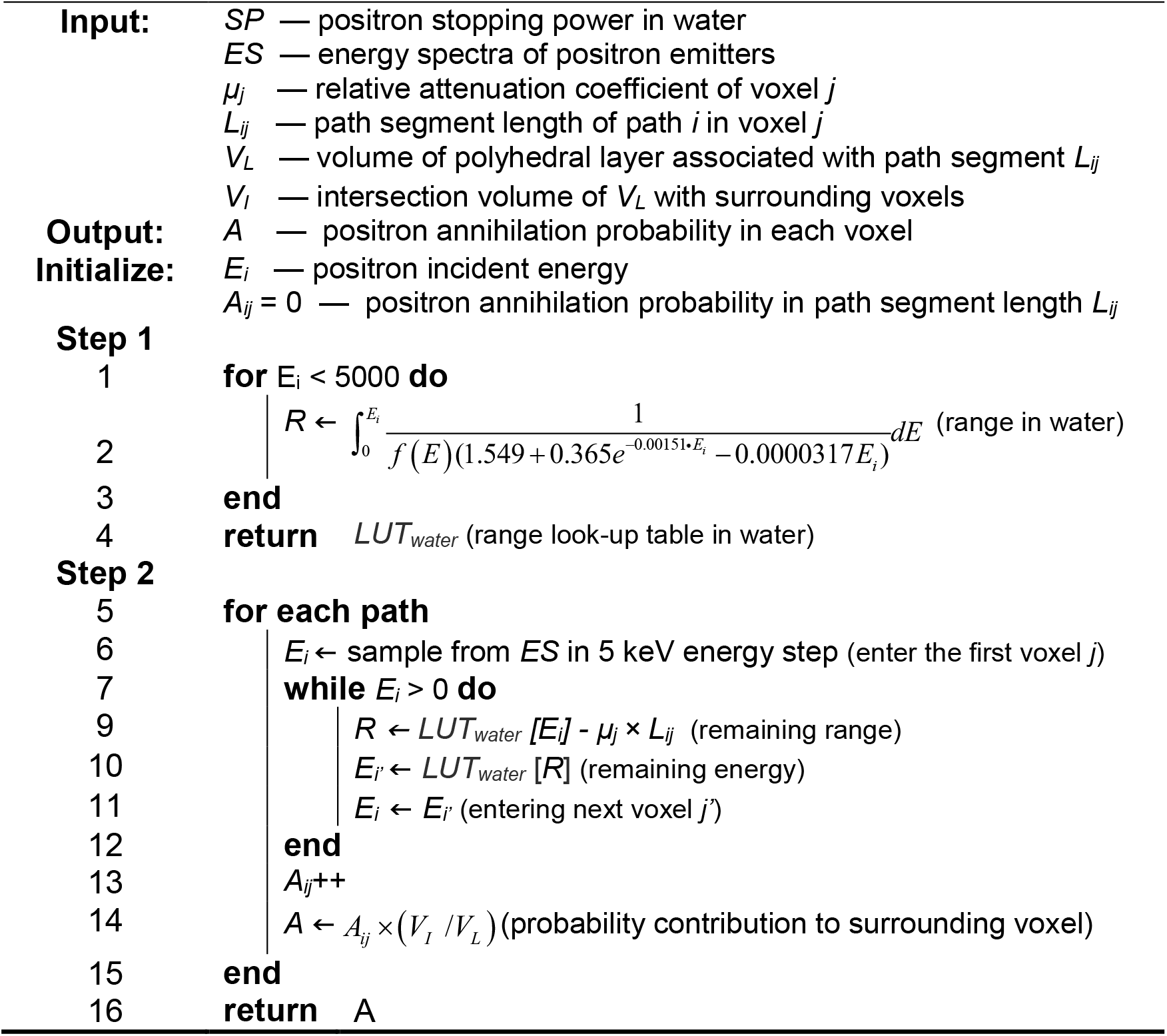

## Notes

### Competing Interest Statement

The authors have declared no competing interest.

